# Peri-Procedural Intravascular Hemolysis during Atrial Fibrillation Ablation: A Comparison of Pulsed-Field with Radiofrequency Ablation

**DOI:** 10.1101/2024.02.15.24302907

**Authors:** Pavel Osmancik, Barbora Bacova, Dalibor Herman, Marek Hozman, Ivana Fiserova, Sabri Hassouna, Vaclav Melenovsky, Jakub Karch, Jana Vesela, Klara Benesova, Vivek Y. Reddy

**Affiliations:** Dept. of Cardiology, University Hospital Kralovske Vinohrady, Charles University, Prague, Czech Republic; Dept. of Laboratory Hematology, Central Laboratories, University Hospital Kralovske Vinohrady, Prague, Czech Republic; Dept. of Molecular Biology, Charles University, Prague, Czech Republic; Institute of Biostatistics and Analyses, Faculty of Medicine, Masaryk University, Brno, Czech Republic; Helmsley Electrophysiology Center, Mount Sinai Fuster Heart Hospital, Icahn School of Medicine at Mount Sinai, New York, New York, USA

## Abstract

**Background:** Despite the favorable safety profile of pulsed-field (PF) energy during ablation of atrial fibrillation (AF), infrequent cases of renal failure, probably caused by hemolysis, have been recently described. The aim was to analyze hemolysis in patients undergoing pulmonary vein isolation (PVI) with PF ablation (PFA) or radiofrequency ablation (RFA).

**Methods:** In consecutive patients, PVI was performed with PFA or RFA. Blood samples were drawn pre-procedure, immediately post-ablation, and one day post-ablation. The concentration of red blood cell microparticles (RBCµ, fragments of damaged erythrocytes) in blood was assessed using flow cytometry (identified as CD235a and annexin V positive events). Lactate-dehydrogenase (LDH), haptoglobin and indirect bilirubin were measured at baseline and at 24 hours.

**Results:** Seventy patients (age 64.7±10.2, 47% women, 36 [51.4%] paroxysmal AF) were enrolled, 47 patients in the PFA group (22 PVI only, 36.4±5.5 PF applications vs. 25 PVI plus additional ablations, 67.3±12.4 PF applications). Twenty-three patients underwent RFA. Compared to baseline, the RBCµ concentration increased ∼ 12-fold post-PFA, and returned to baseline by 24 h (70.8 Nr/µL; 51.8-102.5 vs. 846.6; 639.2-1,215.5 vs. 59.3; 42.9-86.5 Nr/µL, p<0.001); this increase was greater in PVI-plus compared to PVI-only patients (p=0.007). There was also a significant albeit substantially smaller peri-procedural increase in RBCµ with RFA (77.7; 39.2-92.0 vs. 149.6; 106.6-180.8 vs. 89.0; 61.2-123.4,p<0.001). At 24 h with PFA, the concentration of LDH and indirect bilirubin increased, and of haptoglobin decreased highly-significantly (all p<0.001). Only smaller changes in LDH and haptoglobin concentrations (p=0.03), and no change in bilirubin concentrations were present with RFA.

**Conclusion:** PFA was associated with significant peri-procedural hemolysis. The number of PF applications should be minimized.

## INTRODUCTION

Pulsed-field energy (PF) is a new energy source for pulmonary vein isolation (PVI). It uses high-energy, ultra-short electrical pulses (of micro- or nanosecond duration) to irreversibly increase the permeability (electroporation) of cell membranes, with an important degree of preferentiality to ablate cardiomyocyte. (1) It is the first ablation energy ever used that leads to non-thermal cell death, which is in contrast to cryo-, radiofrequency (RF), or laser ablation, where the mechanism of lesion formation is thermal destruction of myocardial tissue.

The first clinically-tested pentaspline catheter for pulsed-field ablation (PFA) of atrial fibrillation (AF) was approved by European regulatory authorities for clinical practice in 2021. In the first human studies on pulsed-filed ablation for atrial fibrillation, PF energy was demonstrated to be a very fast, effective method for PVI with excellent durability. (2) In the first head-to-head clinical comparison with thermal ablation, PFA was non-inferior to RF ablation (RFA) or cryoablation. (3)

Apart from promising clinical results, the safety profile of PFA in the first clinical studies was excellent. (3) (4) However, as with any new treatment method, rare complications can be discovered later, even after thousands of procedures have been performed. Indeed, in the largest database of PF ablations worldwide (*MANIFEST-17K*, with more than 17,000 patients), several cases of renal failure, presumably caused by hemolysis, and requiring temporary dialysis have been reported. (5) Although the final outcome of these patients was positive, the potential for hemolytic effects due to PFA needs to be addressed. Moreover, in our previous study on the effect of PFA on platelets, coagulation, and inflammation, hemolysis was visible in blood samples after the procedure (but not at the beginning or one day later). (6) Therefore, the aim of this study is to analyze hemolysis during PFA and compare it to hemolysis associated with conventional RFA.

## METHODS

### Trial design

This was a prospective, non-randomized, single-center study to evaluate hemolysis during PFA for AF, and to compare it with hemolysis during RFA. The study was approved by the Ethics Committee of the University Hospital Kralovske Vinohrady and was conducted in accordance with the Declaration of Helsinki. Each participant signed informed content before enrollment. The study was registered on clinicaltrials.gov (NCT 06096428).

### Study participants

Patients with symptomatic paroxysmal or non-paroxysmal AF indicated for a first-ever AF ablation procedure were recruited. Patients in whom spontaneous hemolysis could be present (hematological diseases) were excluded, as well as conditions associated with higher bilirubin concentrations (hepatic disorders). Inclusion criteria were the presence of symptomatic paroxysmal or non-paroxysmal AF, age > 18 years, and signed informed content. The exclusion criteria were heart failure with reduced ejection fraction and LV EF < 30%, untreated chronic obstructive pulmonary disease, history of left atrial ablation, and the presence of any malignant, significant hematological, or chronic hepatic disease. The study was non-randomized. The allocation to the type of ablation (PFA vs. RFA) was performed according to the order on the waiting list. In our center, PFA is performed two days/week due to the anesthesia availability, and RFA on the remaining days. Accordingly, the allocation to the particular procedure was based on the patientś position on the waiting list, unless there was a specific patient preference for a particular ablation method. Patients receiving non-vitamin K anticoagulants (NOACs) were given their last NOAC dose the evening before (apixaban, dabigatran) or in the morning (rivaroxaban) of the day before the procedure. Since renal failure due to hemolysis after PFA has only been described in patients receiving high numbers of PF applications during the procedure, we enrolled patients expected to receive both a standard number of PF applications (paroxysmal AF patients) and those expected to require higher numbers of PF applications (non-paroxysmal AF patients).

### Ablation procedures

All procedures were performed under analgosedation using sufentanil, midazolam, propofol, ramimazolam, and ketamine; sedation was milder in RFA patients and deeper in PFA patients. Femoral venous access was achieved using ultrasound guidance. In RF patients, two sheaths (6-Fr, 11-Fr) were inserted in the left femoral vein, one for a 10-Fr phased-array intracardiac echocardiography (ICE) probe (AcuNav, Siemens, Erlangen, Germany) and the other for a decapolar catheter inserted into the coronary sinus (Dynamic XT Catheter, Boston Scientific, MA, USA). In PFA patients, the left femoral vein was used for the decapolar coronary sinus (CS) catheter only in patients for whom a cavotricuspid or mitral isthmus ablation was planned; otherwise, the left femoral vein was left untouched. In all patients, two sheaths were inserted in the right femoral vein: one 11-Fr sheath for the ICE and an 8-Fr for transseptal puncture in the PFA group, and two 8-Fr sheaths, both for transseptal puncture, in the RFA group.

Transseptal punctures were performed using a non-steerable transseptal sheath (SL1, Abbott, MN, USA) under ICE guidance. In PFA patients, the SL1 sheath was replaced by a 13-F deflectable transseptal sheath (Faradrive, Boston Scientific, Inc.) using the over-the-wire technique. Peri-procedural anticoagulation was managed using heparin at a dose of 5000 IU before the transseptal puncture in both groups; another bolus of 5000–10000 IU was given immediately after the transseptal puncture. Activated clotting time (ACT) was assessed every 10 minutes with a target value of 300 seconds; when this target was achieved, further ACT checks were performed every 20 min.

Patients in the PFA group underwent ablation using a pentaspline catheter (Farawave, Boston Scientific, Inc.) with a PFA generator (Farastar, Boston Scientific, Inc.). In patients with paroxysmal AF, only PVI was performed. Ablations were performed using a biphasic bipolar waveform in the following order: four applications with the ablation catheter in the “basket” configuration and four applications with the ablation catheter in the “flower” configuration for each PV ostium. Then, all PVs were checked for entrance and exit block using a standard circular mapping catheter (Lasso, Biosense-Webster Inc., Diamond Bar, CA, USA). If entrance or exit block was not present, additional PF applications were placed to the particular vein. In patients with non-paroxysmal fibrillation, PVI was performed in the same manner as in the paroxysmal AF patients, but ablation of the posterior wall and mitral isthmus was also conducted. For the posterior wall, at least three pairs of PF applications were delivered to each overlapping posterior wall location to connect the right superior and the left superior PV and the right inferior and the left inferior PV (LIPV). The mitral isthmus was ablated between the anterior part of the LIPV and the mitral annulus, typically using 3 – 9 PF applications; conduction block across the posterior mitral isthmus was verified by pacing from the left atrial appendage using the pentaspline PFA (or circular mapping) catheter, and examining the CS activation sequence. If block was not present after the first set of PF applications, additional applications were placed to achieve block of the posterior mitral isthmus. In patients with documented typical atrial flutter, a cavotricuspid-isthmus (CTI) line was also created with PFA, with the catheter in either basket or flower configuration. Bidirectional CTI was evaluated by stimulation of CS and pentaspline PFA catheters.

In RFA patients, all procedures were performed using the CARTO 3 mapping system (Biosense-Webster, CA, USA). A circular mapping catheter was inserted in all PVs for verification of entrance and exit blocks. In non-paroxysmal patients, left atrium (LA) mapping was performed using an Octarey Mapping Catheter (Biosense-Webster, CA, USA). A 3.5 mm irrigated-tip catheter (ThermoCool SmartTouch, or QDot, Biosense-Webster, CA, USA) was also used for mapping and ablation. Ablation was performed using an ablation index target value of 400–450 on the anterior and superior aspects of the PVs and 350–400 on the posterior and inferior aspects. The ultra-high-power, short-duration setting (i.e., QDot plus) was not used. In non-paroxysmal patients, additional ablation was at physician discretion. This additional ablation included the ablation of fractionated signals within scar areas in the left atrium (LA), the ablation of complex fractionated signals, and linear ablations. Accordingly, in patients with documented CTI flutter, a CTI line was created using the same RF catheter.

### Blood sampling

All blood samples were obtained while patients were in a fasting state. Three blood samples were drawn: (T1) at the beginning of the procedure (from the first venous femoral access, i.e. F11 sheath), (T2) from the same venous sheath at the end of the procedure before sheath removal, and (T3) during the morning on the day after the ablation (antecubital vein). In all three samples, the first 5 mL of blood was discarded. Samples from antecubital veins were drawn without tourniquets. Flow cytometry, biochemistry, and hematology were done immediately or within 3 hours of collection.

### Flow Cytometry of Red Blood Cell Microparticles

Samples of citrated whole blood were used to measure the concentration of red blood cell microparticles (RBCµ). Firstly, the platelet-free plasma (PFP) was prepared using two successive 15-min centrigugations at 2,500 g. After the second centrifugation, 100 µL of purified PFP was incubated with 5µL of PE-conjugated CD235a (glycophorin A) and 1µL of FITC-conjugated Annexin V for 20 minutes in the dark. After incubation, 400 µL of ice-cold 1X Annexin V binding buffer and 100 µL of Flow Count Fluorospheres were added to each specimen and mixed gently. All reagents were purchased from Beckman Coulter, Brea, California, USA. The prepared specimens were analyzed within 30 minutes using a Navios EX Flow Cytometer (Beckman Coulter, Brea, California, USA) with the flow rate set at low.

Forward scatter channel (FSC), side scatter channel (SSC) and channels for fluorescent parameters were set to logarithmic gain. The discriminator was set on the FL1 channel with the value 1. For clear resolution of RBCµ from noise, the FSC collection angle was set to the enhanced wide angle (W2) mode. Megamix-Plus FSC^TM^ calibration fluorescent beads with sizes of 0.1 µm, 0.3µm, 0.5 µm and 0.9µm (BioCytex, Marseille, France) were used to demarcate the RBCµ analysis region. RBCµ were therefore identified as CD235a^+^ and Annexin V^+^ events with light scatter distribution within the 0.5 µm and 0.9µm bead range. (**Supplementary Figure 1**) The acquisition stop condition for each sample was set to achieve 2000 events in the flow-count fluorospheres gate.

Collected data were analyzed using Kaluza C version 1.1 software. Proper identification of RBCµ was confirmed by back-gating of CD235a^+^ and Annexin V^+^ events relative to the distribution of calibration beads in the PE x FSC, and SSC x FSC scattergrams. (**Supplementary Figure 1**)

The absolute number of RBCµ per µL of plasma was calculated using the equation provided in the manufacturer’s manual for Flow Count Fluorospheres. Assayed concentrations of Flow Count Fluorospheres was defined as 1,014/µL.

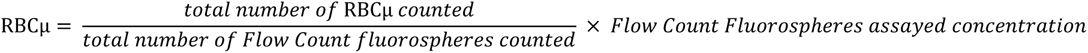

### Biochemistry analysis

Lactate dehydrogenase (LDH), haptoglobin, total bilirubin, and direct bilirubin were quantitatively measured in our clinical biochemistry laboratory. LDH was measured using commercial Atellica CH Lactate Dehydrogenase L-P (LDLP) absorption spectrophotometry assays using Siemens Atellica Solution Analyzer (Siemens Healthineers, Erlangen, Germany). Haptoglobin was measured using commercial N Antiserum to Human Haptoglobin immunonephelometric assay in Atellica NEPH 630 System (Siemens Healthineers, Erlangen, Germany). Total bilirubin and conjugated (direct) bilirubin were quantitatively measured using commercial Atellica CH Total Bilirubin_2 (TBil_2) and Atellica CH Direct Bilirubin 2 (DBil_2) absorption spectrophotometry assays in a Siemens Atellica Solution Analyzer (Siemens Healthineers, Erlangen, Germany). Values of total bilirubin and conjugated bilirubin were used for the calculation of unconjugated (indirect) bilirubin.

### Complete blood count and enumeration of schistocytes

Complete blood counts with differential and reticulocytes were measured in our clinical hematology laboratory. Samples of whole blood collected in EDTA blood collection tubes were analyzed using automatic Sysmex XN 1000 analyzer (Sysmex Corporation, Kobe, Japan).

### Statistical analysis

Standard descriptive statistics were used in our analyses. Binary or categorical parameters of patients were characterized by absolute and relative frequencies, while continuous parameters were described as means and standard deviations. Since most markers did not have a normal distribution, the median and interquartile ranges (IQR) were used to describe those parameters. The Mann-Whitney test was used to assess the differences in continuous parameters between groups (i.e., PFA vs. RFA, PVI-plus vs. PVI-only), and the Fisheŕ exact test was used for categorical parameters. In related samples, Friedman’s two-way analysis of variance (ANOVA) by ranks with a Bonferroni correction for post hoc testing was employed for evaluating the progression of individual markers. However, when only baseline (T1) and discharge (T3) marker levels were assessed, the related samples Wilcoxon signed rank test was utilized. Spearman’s correlation coefficient was calculated to test the association between variables. The level of statistical significance for all analyses was p = 0.05. Analyses were performed in SPSS 28.0.1.1 (IBM Corporation, Armonk, NY, USA, 2021).

### Sample size consideration

No data had been published on hemolysis for either radiofrequency or pulsed-field ablation before the study was initiated. Since in most previous papers on intravascular hemolysis during interventional cardiovascular procedures, LDH or haptoglobin were determined, these two markers were initially considered for sample size calculation. Because LDH is also present in cardiomyocytes and its concentrations are elevated during myocardial injury (myocardial infarction, or catheter ablation), the sample size was calculated based on expected changes in haptoglobin concentration. In an absence of inflammation, haptoglobin decrease reflects the degree of hemolysis. The power calculation was assessed based on the expected differences in haptoglobin concentration. In the pilot sample of 12 patients, in whom haptoglobin concentrations were measured before and after pulsed-field ablation, the concentration of haptoglobin decreased by 0.7 with a standard deviation of 0.27. A sample size of at least 22 patients needed to be enrolled in any group to detect a decrease of at least 0.2 mmol/l, standard deviation = 0.27 with a power = 90%, and an alpha = 0.05. To further determine the differences in concentrations between PF patients undergoing only PVI, vs. PVI plus additional ablations, at least 44 PFA patients (22 PVI only and 22 PVI plus) were planned for enrollment. A power calculation was not done for all measured biomarkers. Since haptoglobin is relatively specific for hemolysis, this sample was considered sufficient even for other hemolytic markers.

## RESULTS

Between November 2023, and February 2024, 70 patients (age 64.7±10.2 years, 33 [47%] women, 36 [51.4%] with paroxysmal AF) were enrolled. Baseline clinical characteristics are shown in **Table 1**. The procedure was substantially faster with PFA compared to RFA (57.0±15.3 vs. 144.2±48.5 min, p<0.001), which was also true of the LA dwell time (36.0±13.5 vs 107.5±52.4 min, p<0.001). One minor (small pseudoaneurysm with manual compression without hospitalization prolongation) and no major complications occurred. Procedural characteristics are shown in **Table 2**.

**Table 1.**
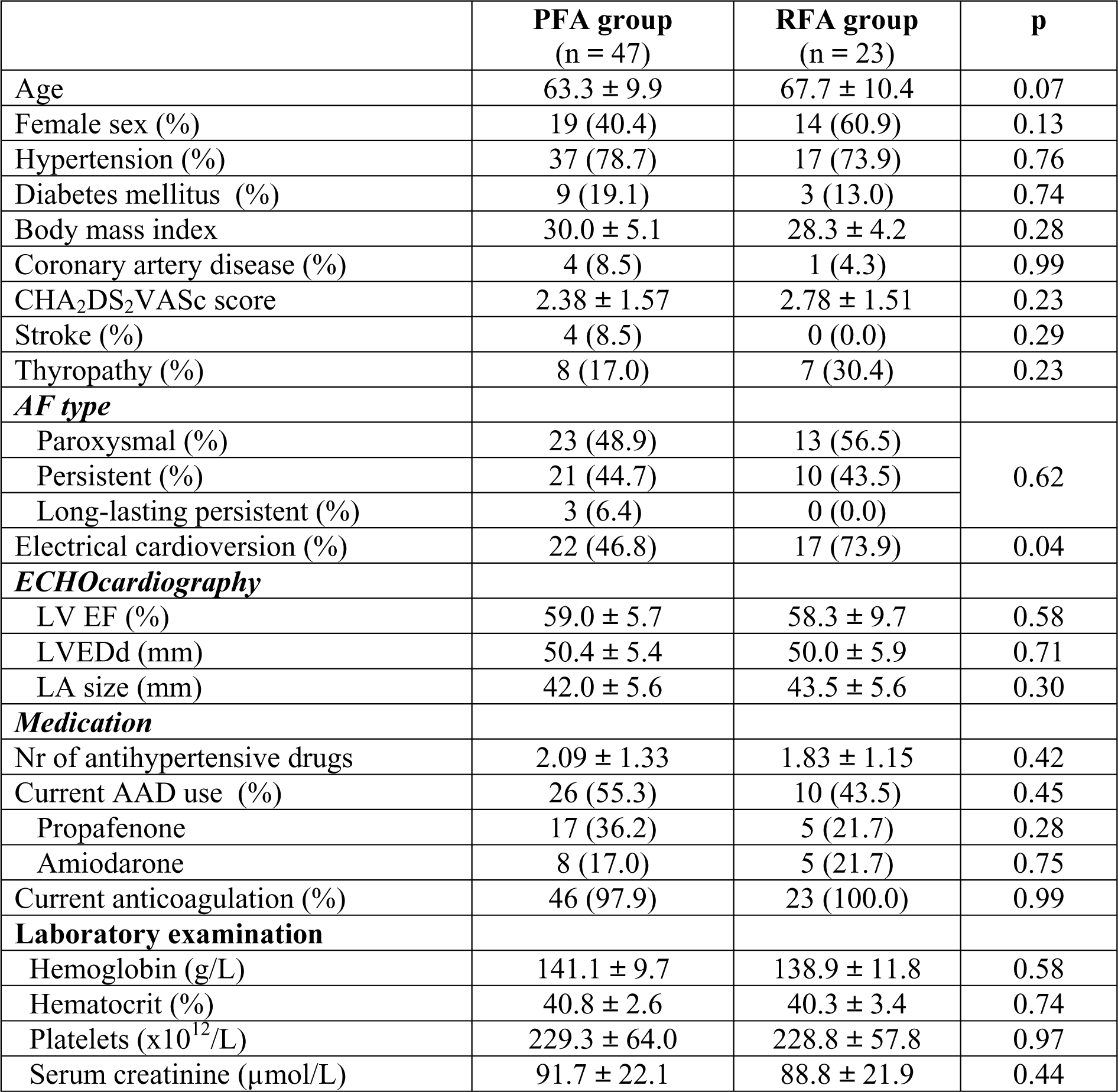
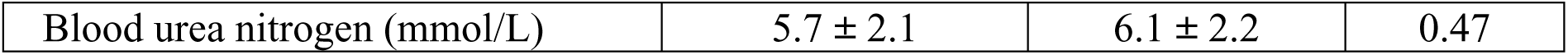
Baseline characteristics. LV EF – left ventricular ejection fraction, LVEDd – left ventricular end-diastolic dimension, LA – left atrium, AAD – antiarrhythmic drug

**Table 2.** Procedural characteristics. LA- left atrium, IU – international unit, ACT – activated clotting time, PFA – pulsed-field ablation, RF – radiofrequency, PV – pulmonary vein, CTI – cavotricuspid isthmus

|  | **PFA group**<br>(n = 47) | **RFA group**<br>(n = 23) | **P** |
| --- | --- | --- | --- |
| Procedure duration (min) | 57.0 ± 15.3 | 144.2 ± 48.6 | < 0.001 |
| Fluoroscopy duration (min) | 7.1 ± 3.1 | 5.6 ± 3.4 | 0.01 |
| LA dwelling time | 36.0 ± 13.6 | 108.0 ± 52.4 | < 0.001 |
| Total heparin dose (IU) | 15 383 ± 1 714 | 13 826 ± 4 097 | 0.05 |
| Maximum ACT (sec) | 360.0 ± 50.8 | 361.9 ± 65.5 | 0.51 |
| Electrical cardioversion (%) | 21 (44.7) | 10 (43.5) | 0.99 |
| SR at discharge (%) | 46 (97.9) | 22 (95.7) | 0.99 |
| ***Ablation*** |  |  |  |
| Total No of LA PFA applications (No) / total LA RF ablation time (min) | 52.9 ± 18.4 | 29.1 ± 9.6 | NA |
| No of PFA applications on PVs (No) / PV RF ablation time (min) | 34.9 ± 4.2 | 22.9 ± 7.9 | NA |
| No of patients with CTI ablation (%) | 4 (8.5) | 4 (17.4) | 0.42 |

Based on the type of AF and the scheduled procedure, PFA patients were further divided into PVI-only and PVI-plus subgroups. The clinical and procedural characteristics of these two subgroups are shown in **Table 3**. The PVI-plus approach was more often used in non-paroxysmal AF patients (92% vs 4.6%) and as expected, the procedures were of longer duration (63.4±13.5 vs. 49.7±14.2, p=0.001). PVI-plus patients received significantly more PF applications (67.3±12.4 vs. 36.4±5.5, p<0.001).

**Table 3.**
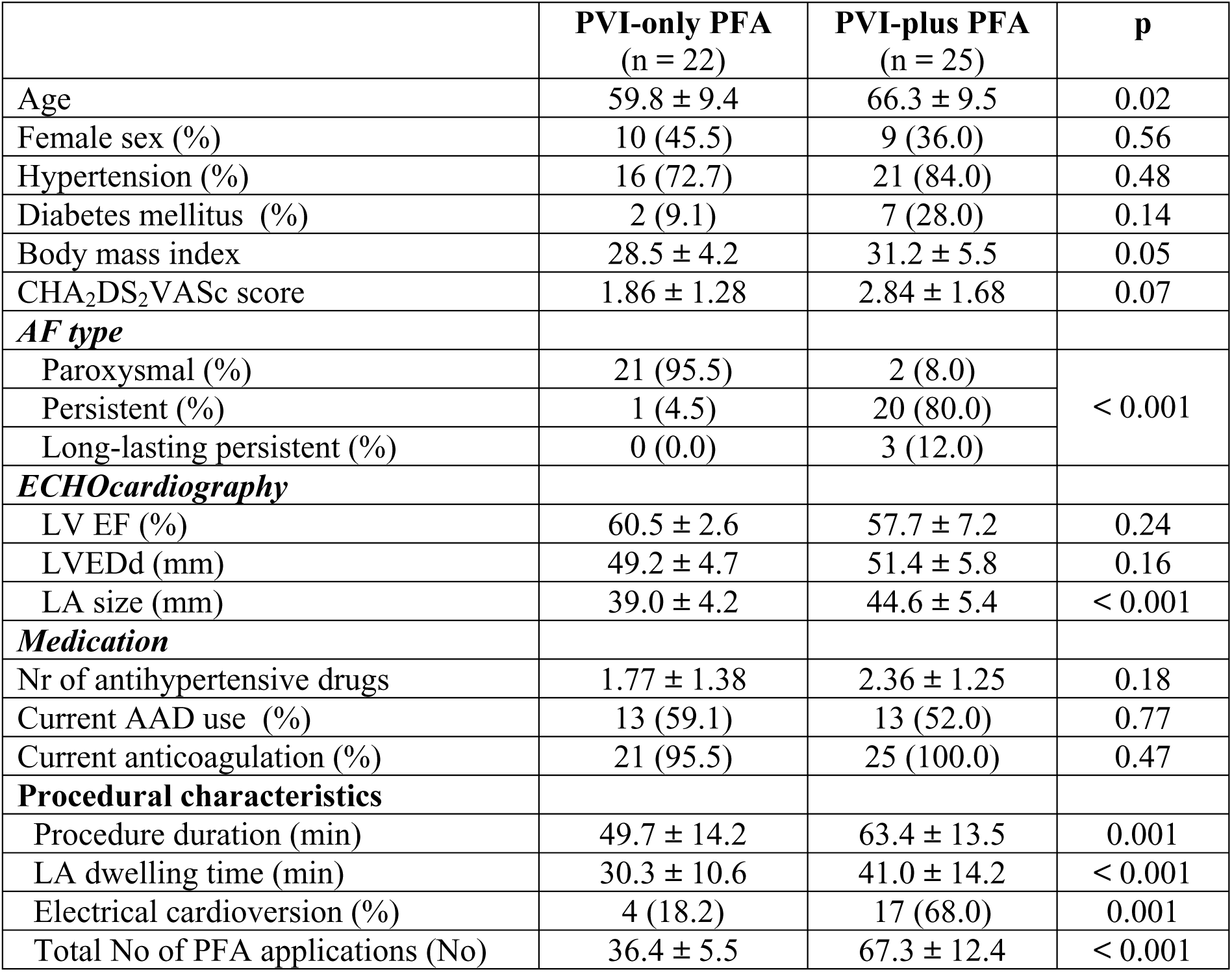
Baseline characteristics and differences between PVI-only and PVI-plus patients. LV EF – left ventricular ejection fraction, LVEDd – left ventricular end-diastolic dimension, LA – left atrium, AAD – antiarrhythmic drug, PFA – pulsed-field ablation

|  | **PVI-only PFA**<br>(n = 22) | **PVI-plus PFA**<br>(n = 25) | **p** |
| --- | --- | --- | --- |
| Age | 59.8 ± 9.4 | 66.3 ± 9.5 | 0.02 |
| Female sex (%) | 10 (45.5) | 9 (36.0) | 0.56 |
| Hypertension (%) | 16 (72.7) | 21 (84.0) | 0.48 |
| Diabetes mellitus (%) | 2 (9.1) | 7 (28.0) | 0.14 |
| Body mass index | 28.5 ± 4.2 | 31.2 ± 5.5 | 0.05 |
| CHA<sub>2</sub>DS<sub>2</sub>VASc score | 1.86 ± 1.28 | 2.84 ± 1.68 | 0.07 |
| ***AF type*** |  |  |  |
| Paroxysmal (%) | 21 (95.5) | 2 (8.0) | < 0.001 |
| Persistent (%) | 1 (4.5) | 20 (80.0) |  |
| Long-lasting persistent (%) | 0 (0.0) | 3 (12.0) |  |
| ***ECHOCardiography*** |  |  |  |
| LV EF (%) | 60.5 ± 2.6 | 57.7 ± 7.2 | 0.24 |
| LVEDd (mm) | 49.2 ± 4.7 | 51.4 ± 5.8 | 0.16 |
| LA size (mm) | 39.0 ± 4.2 | 44.6 ± 5.4 | < 0.001 |
| ***Medication*** |  |  |  |
| Nr of antihypertensive drugs | 1.77 ± 1.38 | 2.36 ± 1.25 | 0.18 |
| Current AAD use (%) | 13 (59.1) | 13 (52.0) | 0.77 |
| Current anticoagulation (%) | 21 (95.5) | 25 (100.0) | 0.47 |
| ***Procedural characteristics*** |  |  |  |
| Procedure duration (min) | 49.7 ± 14.2 | 63.4 ± 13.5 | 0.001 |
| LA dwelling time (min) | 30.3 ± 10.6 | 41.0 ± 14.2 | < 0.001 |
| Electrical cardioversion (%) | 4 (18.2) | 17 (68.0) | 0.001 |
| Total No of PFA applications (No) | 36.4 ± 5.5 | 67.3 ± 12.4 | < 0.001 |

### Comparison between PFA and RFA patients

In total, 47 patients underwent PF ablation and 23 RF ablation. The markers of hemolysis were significantly increased with PFA compared to RFA (**Central illustration**). Moreover, hemolysis was higher in the PVI-plus patients compared to PVI-only patients (**Central illustration**). The time-course of concentrations of RBCµ is shown in **Figure 1**. The concentration of RBCµ at the end of the procedure compared to baseline value was significantly increased in both groups (**Figure 1**); however, this increase was significantly higher in the PFA group compared to the RFA group (p<0.001). Compared to baseline values, the peak RBCµ in PFA patients was ∼12 times higher than baseline values (in contrast, this increase was ∼ two-fold in RFA patients). In both groups, the concentration of RBCµ had returned to baseline values by the day after the procedure.

**Figure 1.**
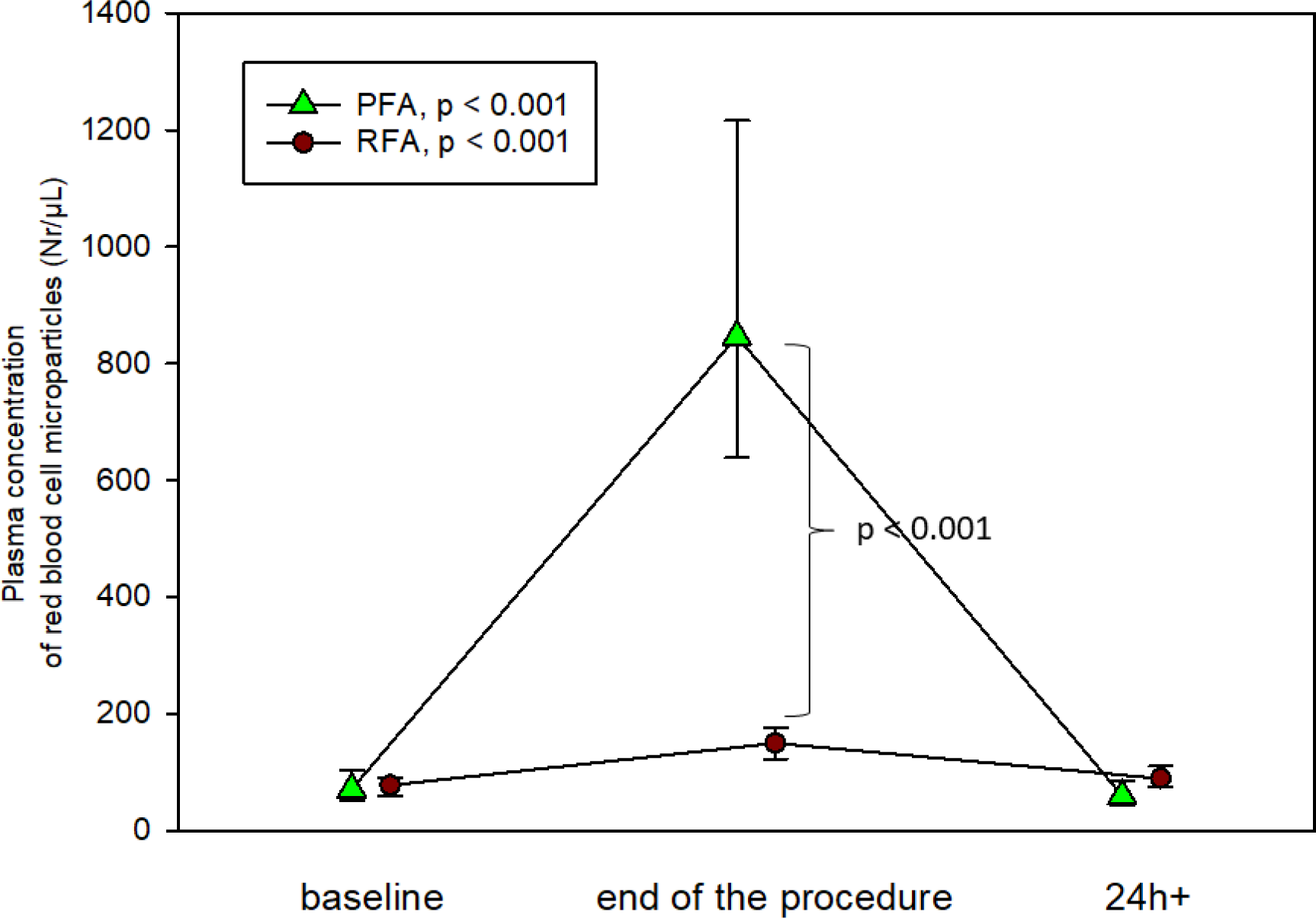
The time course of plasma concentrations of red blood cell microparticles during PF and RF ablation. Figures display medians with IQRs (PFA – green triangles, RFA – red circles). Progression results for each group are shown in rectangles. Significant differences between both groups are shown for specific times.

The concentration of LDH (2.70; 2.38-2.91 vs. 4.67; 4.05-5.46, p<0.001) and indirect bilirubin (10.3; 7.5-12.8 vs. 14.1; 10.1-18.7, p<0.001) significantly increased, while haptoglobin significantly decreased (1.20; 0.87-1.48 vs. 0.44; 0.25-0.71, p<0.001) on the day after the procedure in the PFA patients (**Figure 2 & 3**). In the RFA patients, concentrations of LDH (2.89; 2.53-3.25 vs. 3.08, p=0.027) increased and haptoglobin (1.31; 0.92-1.46 vs. 1.17; 0.94-1.44, p=0.03) decreased significantly one day after the procedure (**Figure 2 & 3**), but this difference was substantially smaller compared to the PFA group. No significant increase in indirect bilirubin (10.1; 7.4-14.8 vs. 9.3; 7.8-15.8, p=0.54) concentrations were present in the RFA patients. No changes in reticulocytes or the immature fraction of reticulocytes were present in PFA or RFA patients (**Figure 3**).

**Figure 2.**
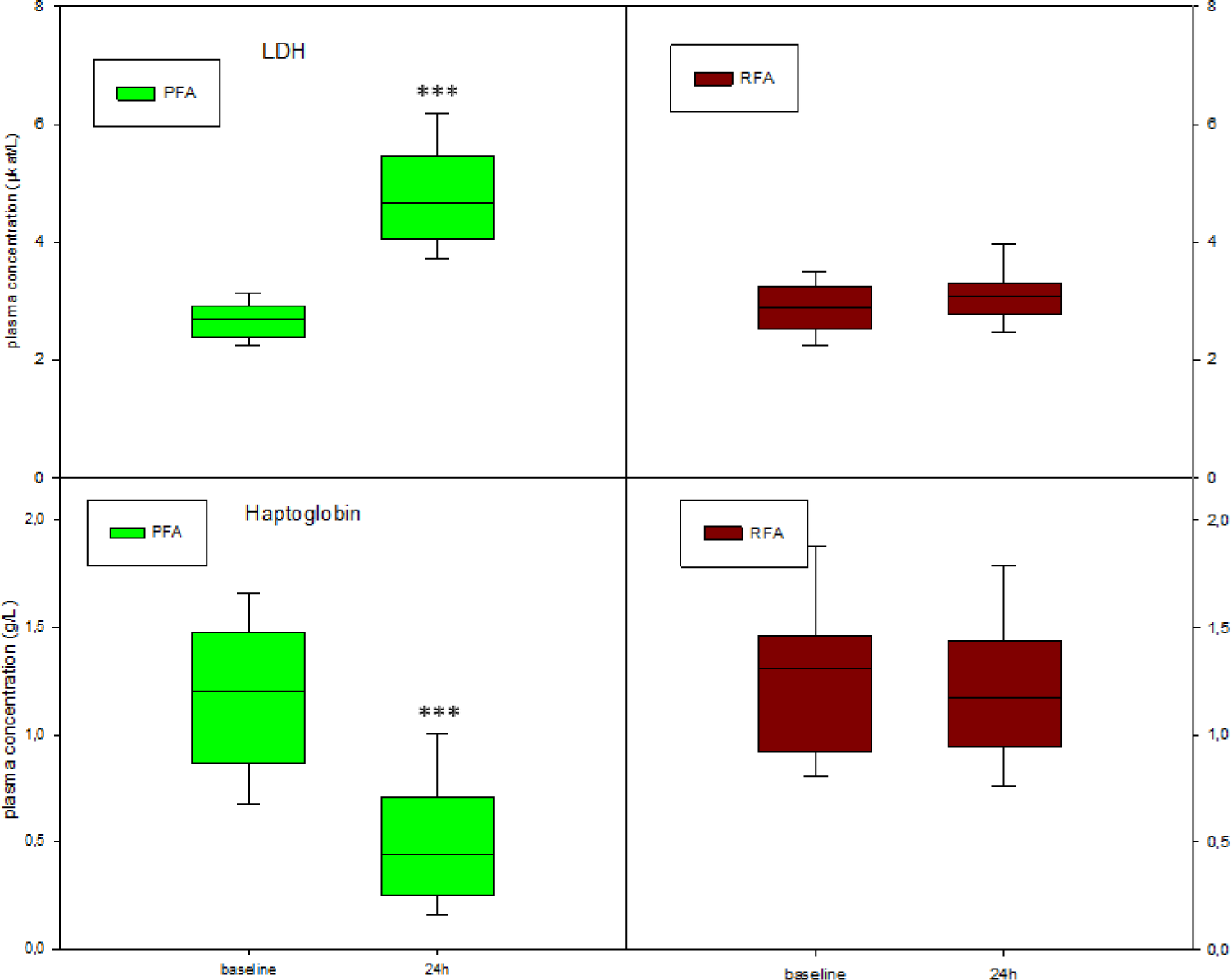
The concentrations of lactate-dehydrogenase and haptoglobin before and 24 h after ablations. Box plots display the medians with IQRs (PFA – green rectangles, RFA – red rectangles). Significant differences between both groups are shown for specific times (*** p<0.001).

**Figure 3.**
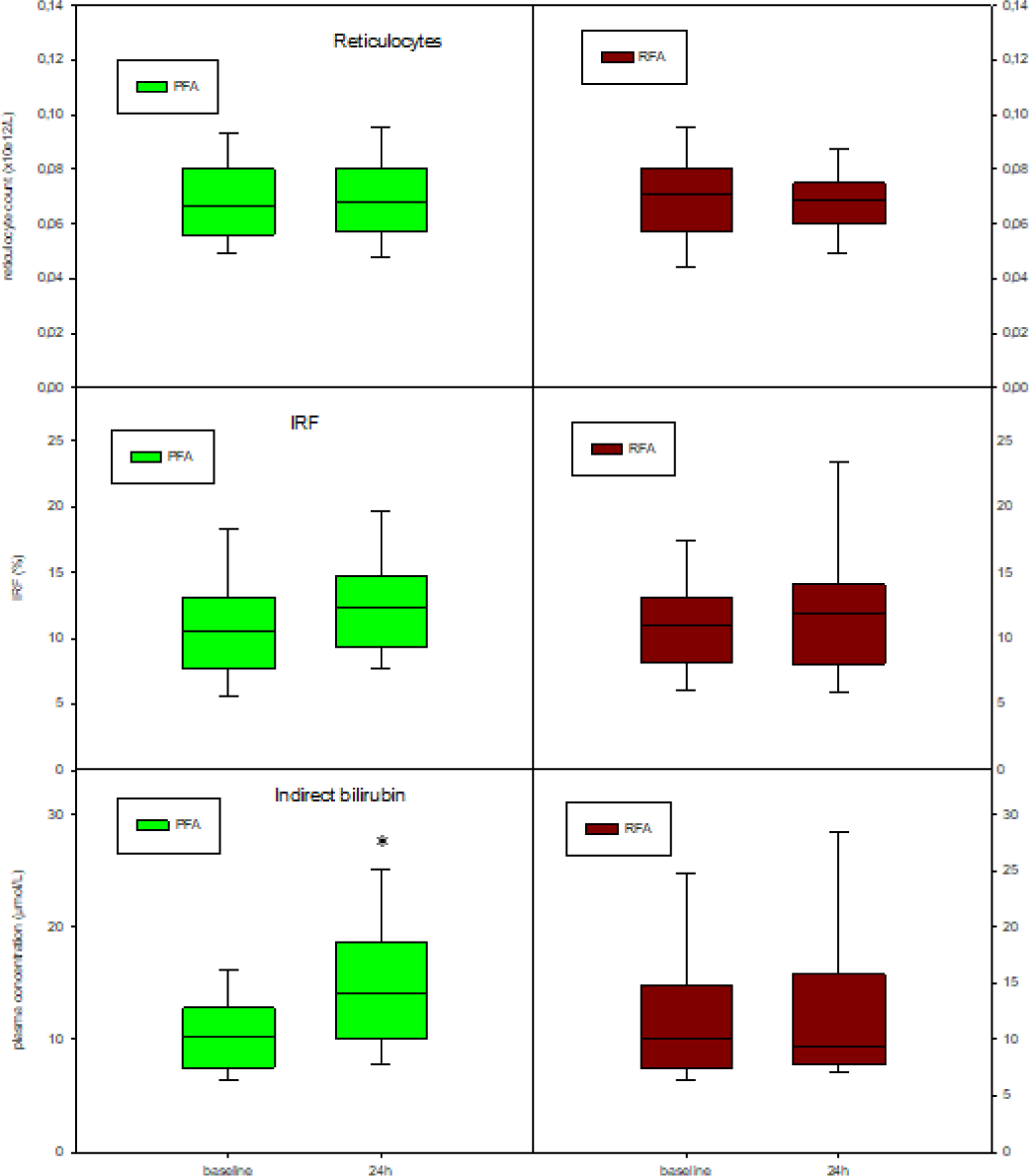
Concentrations of reticulocytes, the immature fraction of reticulocytes, and indirect bilirubin before and 24 h after ablations. Box plots display medians with IQRs (PFA – green rectangles, RFA – red rectangles). Significant differences between both groups are shown for specific times (* p<0.05).

### Comparison between PVI-only and PVI-plus subgroups among the PFA patients

In the PFA group, 25 patients underwent PVI plus additional ablations (posterior wall and mistral isthmus), while 22 patients underwent PVI only. Procedures, as well as the LA dwelling times, were longer in PVI-plus patients (**Table 2**). Baseline values of all measured biomarkers were similar between both subgroups. The peak concentration of RBCµ was significantly higher (657.1; 502.3-1,006.5 vs. 924.2; 758.6-1,355.5, p=0.007) in PVI-plus vs. PVI-only patients (**Figure 4**). Similarly, the concentration of LDH on the day after the ablation was significantly higher (4.21; 3.80-4.67 vs. 5.24; 4.44-5.88, p<0.001), and the concentration of haptoglobin was significantly lower in the PVI-plus patients (0.62; 0.34-0.88 vs. 0.35; 0.20-0.59, p=0.04). However; the concentrations of indirect bilirubin did not differ between groups (12.5; 8.9-15.6 vs. 17.4; 11.8-19.1, p=0.08) (**Supplementary Figure S2**).

**Figure 4.**
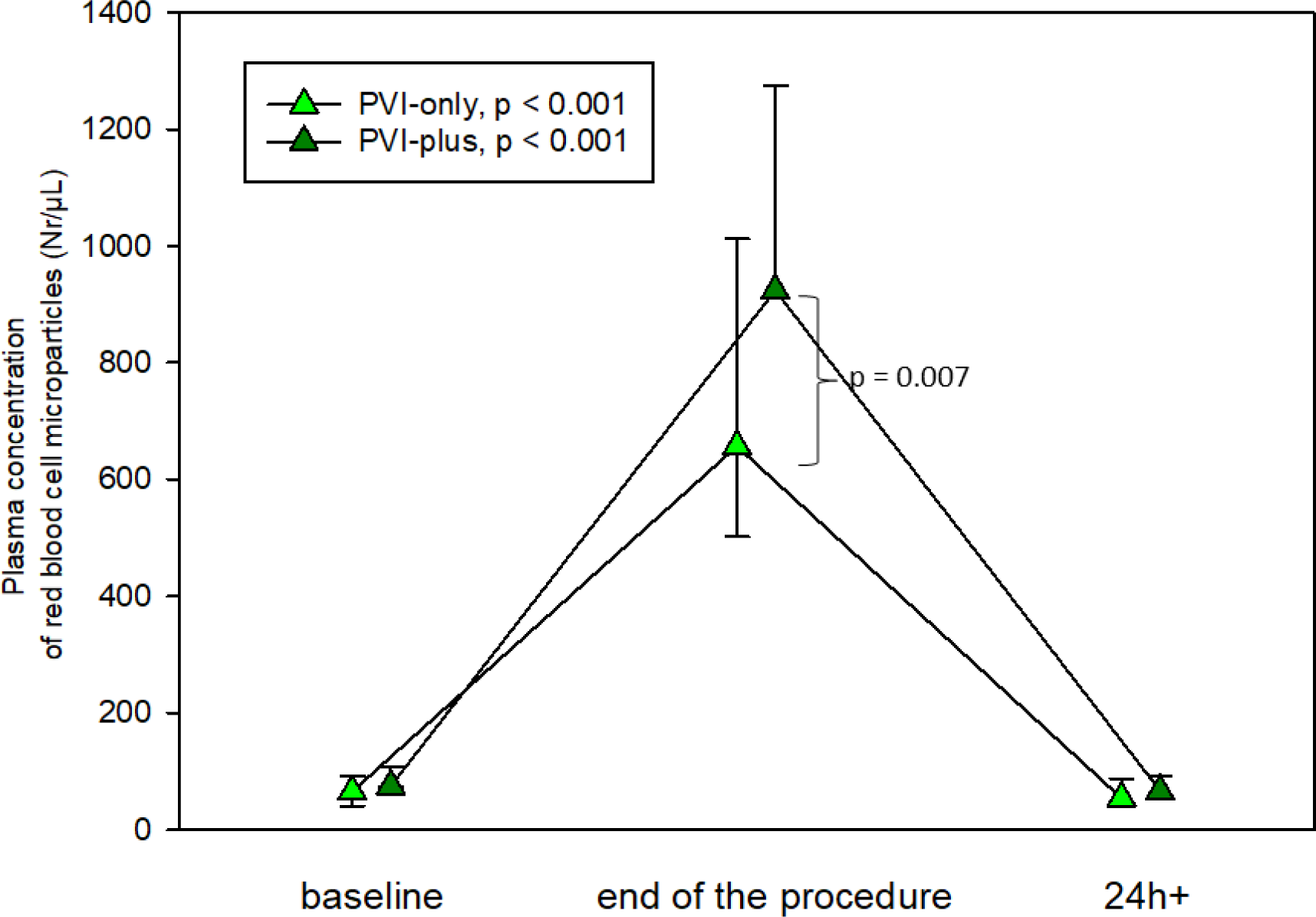
The time course of concentration of red blood cell microparticles in PVI-only vs. PVI-plus patients. Figures display medians with IQRs (PVI-only: light green triangles, PVI-plus: dark green triangles) Progression results for each group are shown in rectangles. Significant differences between both groups are shown for specific times.

There were no differences in reticulocytes, or the immature fraction of reticulocytes between PVI-plus and PVI-only patients (**Supplementary Figure S3**).

### Correlation between the number of impulses and markers of hemolysis

A positive correlation was found between the total number of applications with the concentration of RBC microparticles at the end of ablation (r=0.31, p=0.03), and also with the concentration of LDH one day after the procedure (0.4, p=0.006). There was an inverse, but not significant correlation between the total number of applications with the plasma haptoglobin level (p=0.09).

## DISCUSSION

In our study of patients with pulsed-field vs. radiofrequency ablation, a significant amount of hemolysis was observed during PFA. Furthermore, the amount of hemolysis increased with the number of PF applications.

### Hemolysis during cardiovascular procedures

Mechanical intravascular hemolysis occurs when red blood cells in blood vessels are fragmented by mechanical or other injury. It is one of the most frequently encountered problems during mechanical circulatory support. (7) The reported incidence of hemolysis during the use of percutaneous ventricular assist using microaxial flow pump (Impella) reaches 32%. (8) Heart valve replacement with mechanical prosthesis, even with new-generation prosthetic heart valves, is also associated with mild intravascular hemolysis. (9) Significant mechanical hemolysis related to secondary mechanical destruction of red blood cells passing through paravalvular leaks has also been reported. (10) The mechanism of hemolysis with a ventricular assist device, or mechanical valves, differs from the mechanism of hemolysis linked to PFA. Hemolysis during ablation is not caused by mechanical damage to RBCs but is more likely caused by the direct effect of the high-voltage energy current on RBCs.

### Hemolysis and electrical field

In pre-clinical studies, high cardiac selectivity was attributed to PFA with no or low damage to adjacent structures. Pre-clinical studies on PFA evaluated damage to the phrenic nerve, coronary arteries, esophagus, aorta, and closely adjacent structures such as ganglionated plexi or Purkinje fibers. (11) Surprisingly, no current report (either *in vitro* or *in vivo*) specifically addresses damage to erythrocytes during RFA or PFA, although the PF catheter is in direct contact with the blood during every ablation.

Publications on the effect of PFA on erythrocytes are sparse. In the 1990s, the local deformation of RBCs in high-frequency electric fields was studied. In *in vitro* experiments, RBCs were exposed to alternating electric fields of 1 MHz frequency and 0 - 4000 V/cm intensity by means of two microelectrodes placed at a distance of 10 - 20 µm from the cell surface. The direct deformation process on RBC membranes involved pulling the cell membranes toward the electrodes, which was observed using a phase contrast microscope, and the deformation was proportional to the square of the field intensity. (12) Importantly, increasing the field intensity to 5000 - 5500 V/cm led to swelling and hemolysis of a considerable number of RBCs. (12) As noted, information on the *in vitro* effect of the exact PF pulse on erythrocytes that is used during *in vivo* catheter ablation is lacking. Significant hemolysis leading to various degrees of hemoglobinuria, hemolytic jaundice or even acute kidney injury has been repeatedly described in patients with giant hepatic hemangioma treated by radiofrequency ablation, which documents the in vivo effect of electric field (although of different energy) on the erythrocytes. (13)

Theoretically, intravascular hemolysis can also be a consequence of thrombosis or activation of coagulation due to an exposure to foreign material. However, no excess in clinical thrombotic events during PFA has been reported in large PFA studies. (4) Also, as shown recently, platelet activation and coagulation changes are very similar during RFA and PFA. (6) Therefore, the cause of hemolysis is more liked to the direct effect of electric fields on erythrocytes than to thrombosis. No changes in reticulocyte count was present one day after ablation, but it could be expected later on or during repeated attacks of hemolysis.

### Clinical consequence of hemolysis

In general, the clinical consequence of hemolysis depends on the acute intensity of hemolysis, its duration, and the volume status of the patient. In patients with a mechanical valvular prothesis, typically only mild elevation of hemolysis markers and mild anemia is present. (14) Renal impairment occurs only in the presence of massive and rapid intravascular hemolysis, especially in hypovolemic patients with pre-existing renal disease. Plasma-free hemoglobin that is delivered after RBC damage is bound to haptoglobin in order to prevent iron loss and hemoglobin-mediated renal injury. There is sufficient haptoglobin in circulation to bind and clear 3 g of hemoglobin. When the binding capacity of serum haptoglobin is fully saturated, the concentration of plasma-free hemoglobin increases, and the accumulation of hemoglobin derivatives in the proximal tubule cells could lead to acute tubular necrosis. In a large series of 17,000 patients in the *MANIFEST-17K* study recently reported on American Heart Association, renal failure requiring dialysis occurred in 5 (0.03%) patients. (5) In the very recent report by Venier et al, two patients experienced acute kidney injury with hospitalization prolongation after PFA for AF. (15)

In our study, a clinically-relevant increase of creatinine (to 181 µmol/L) was present in only one patient. This patient received 65 PF applications, but his pre-procedural creatinine was already elevated (130 µmol/L). Hospitalization was not prolonged, and only increased hydration and repeated laboratory check after discharge was required; further follow-up was uncomplicated with decline of the creatinine concentration to baseline in 10 days. Importantly, as shown in our patients, the extent of hemolysis as determined by RBCµ concentration was higher in patients with more PF applications. In the *MANIFEST-17K* study, patients who required temporary dialysis due to acute kidney injury received 143±27 PF applications during the procedure. In the recent report by Venier et al., the two patients who developed acute kidney injury after PFA received 174 and 126 PF applications, respectively. All of these patients received substantially more PF applications than in our series. The average number of PF application in our PVI-plus subgroup was 67.3±12.4, and maximum number was 100 applications (in only one patient). On the other hand, the increase of RBCµ concentration was higher in patients with a higher number of PF applications. Moreover, the highest creatinine value could be expected 2∼3 days after procedure, and therefore, these patients could have been missed. The vast majority of hemolysis was likely subclinical, with no impact on kidney function, and as such, significant kidney injury due to hemolysis during PFA is only the tip of the iceberg.

Our study was not designed for, and is thus incapable of determining the safe threshold for the number of PF applications. However, it seems prudent that a large number of PF applications, especially in patients with pre-existing renal disease, should be avoided.

### Hemolysis during radiofrequency ablation

In our patients, peri-procedural laboratory-detected hemolysis was also present in RFA patients. However; the amount of hemolysis was very small; there was only borderline change in LDH and haptoglobin concentrations (p=0.03). The almost two-fold increase in concentration of RBCµ was more significant, but it is a very sensitive parameter for hemolysis. Of course, hemolytic renal failure has never been described after RFA of cardiac arrhythmias after hundreds of thousands of procedures. Therefore, no report has still addressed the hemolysis during RFA. So while interesting, whatever small extent of hemolysis that may exist with RFA does not seem to be associated with any clinically relevant complications. Small increase in hemolysis with RFA could be associated with the exposure to foreign, non-phospholipid surface of catheter and sheath material, activation of coagulation, or direct effect of electrical field.

On the other hand, clinically-relevant hemolysis with acute kidney injury after radiofrequency ablation has already been described after RF application of giant hemangiomas. Although this clinical scenario differs significantly, it documents the possible effect of the electric field on erythrocytes. (13) In our patients in whom RFA was used for cardiac ablation, the hemolysis was detected primarily as an increased concentration of RBCµ, which as noted above, is very sensitive marker of erythrocyte damage. Currently, there is no standard definition for intravascular hemolysis during cardiovascular procedures, raising the possibility that the extent of RBC damage that we noted might be considered trivial relative to meaningful laboratory hemolysis.

### Markers and hemolysis

Currently, a uniformly accepted definition of intravascular hemolysis based on laboratory parameters is lacking. RBCs are rich in LDH, and fragmentation of RBCs leads to a rapid increase in LDH. However, LDH is a nonspecific marker, since any cause of cellular lysis or necrosis, such as myocardial infarction or ablation injury present during catheter ablation, leads to increased LDH. Haptoglobin is an acute-phase reactant whose major role is binding free plasma hemoglobin. Therefore, with hemolysis, plasma haptoglobin concentration is substantially decreased. As an acute phase reactant, haptoglobin is also decreased during acute inflammatory processes. However, in elective patients without acute inflammation, e.g. patients admitted for elective catheter procedure, decreased haptoglobin, especially in association with increased LDH, becomes very specific for hemolysis. In the report by Vernier et al., a decrease of haptoglobin less than 0.04 g/L was assessed as sign of hemolysis. However, hemolysis is not a binary process. As shown in our patients, the number of RBC fragments presents a continuous variable. Moreover, if haptoglobin capacity is exhausted (after damage of 3 g of hemoglobin), hemolysis may continue but the concentration of haptoglobin cannot follow it. The presence of RBCµ represents a very specific, sensitive and accurate marker of ongoing intravascular hemolysis. As shown in our study, there is ∼ 12-fold increase of RBC microparticles immediately after ablation, which then rapidly returns to pre-procedural values. In contrast to the haptoglobin decrease, this rapid change of the concentration of this marker best reflects the ongoing hemolytic process. Our study was not able to characterize the “safe” number of PF applications, i.e., the number of PF applications associated with renal impairment. However, it was shown that RBCµ concentrations increased with higher number of PF pulses, and therefore, high numbers of PF applications cannot be recommended and should be avoided, especially in patients with pre-existing renal disorders.

## STUDY LIMITATION

The study was non-randomized. Therefore, selection bias cannot be excluded. Mid- or long-term outcomes with regard to renal function, especially tubular necrosis caused by increased hemolysis, were not determined. The study was not able to determine the safety threshold for PF applications.

## CONCLUSION

Hemolysis occurred during pulsed-field ablation of AF, far exceeding what occurs during radiofrequency ablation. The extent of hemolysis depends on the number of PF applications. The numbers of PF applications should be minimized.

## Data Availability

Data will be available upon reasonable request when the manuscript is published

## LIST OF ABBREVATIATION

PFA: pulsed-field ablation
PVI: pulmonary vein isolation
RFA: radiofrequency ablation
AF: atrial fibrillation
NOAC: non-vitamin K anticoagulants
ICE: intracardiac echocardiography
CS: coronary sinus
Act: activated clotting time
LIPV: left inferior pulmonary vein
LA: left atrium
CTI: cavotricuspid isthmus
ELISA: enzyme-linked immunoassay
RBC: red blood cell
PFP: platelet-free plasma
CD: cluster of differentiation
PE: phycoerythrin
FITC: fluorescein isothiocyanate
FSC: forward scatter
SSC: side scatter
LDH: lactate dehydrogenase
EDTA: ethylenediamine tetraacetic acid
IQR: interquartile ranges
MHz: mega Herz

## Central illustration

Summary of results: Figures display the concentration of red blood cell microparticles in patients undergoing pulmonary vein isolation with (PFA-PVI plus) or without additional left atrial ablations using pulsed-field energy (PFA-PVI only) and in patients undergoing pulmonary vein isolation only, using radiofrequency (RFA) energy. PFA–PVI only: light green triangles, PFA–PVI plus: dark green triangles, RFA: red circles. Significant differences between groups are displayed (*** p<0.001 for differences between groups at 24 hours post-procedure)

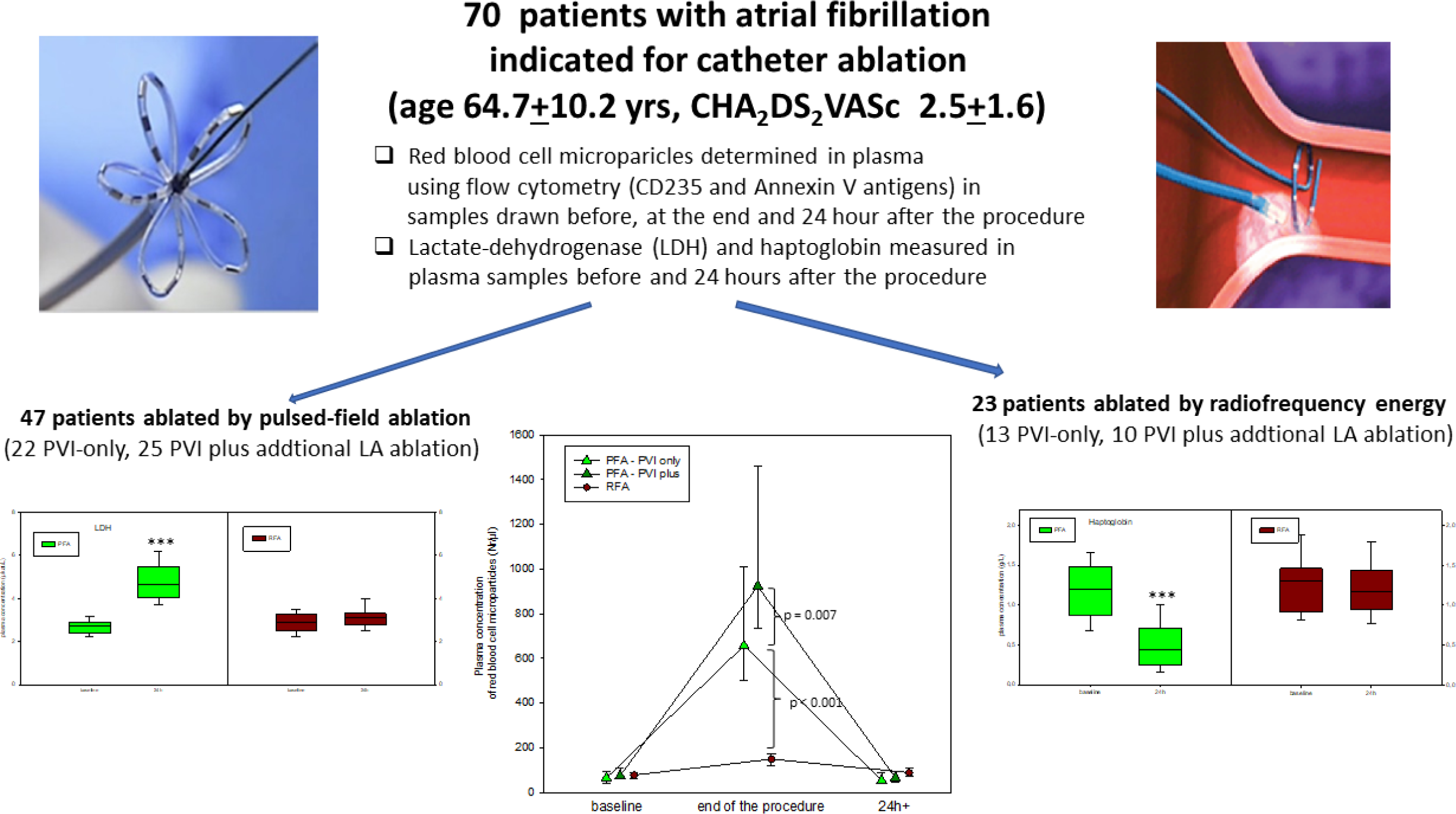

## REFERENCES

1. Grimaldi M, Di Monaco A, Gomez T, Berman D, Datta K, Sharma T, et al. Time Course of Irreversible Electroporation Lesion Development Through Short- and Long-Term Follow-Up in Pulsed-Field Ablation-Treated Hearts. Circ Arrhythm Electrophysiol. 2022;15(7):e010661.

2. Reddy VY, Dukkipati SR, Neuzil P, Anic A, Petru J, Funasako M, et al. Pulsed Field Ablation of Paroxysmal Atrial Fibrillation: 1-Year Outcomes of IMPULSE, PEFCAT, and PEFCAT II. JACC Clin Electrophysiol. 2021;7(5):614–27.

3. Reddy VY, Gerstenfeld EP, Natale A, Whang W, Cuoco FA, Patel C, et al. Pulsed Field or Conventional Thermal Ablation for Paroxysmal Atrial Fibrillation. N Engl J Med. 2023;389(18):1660–71.

4. Ekanem E, Reddy VY, Schmidt B, Reichlin T, Neven K, Metzner A, et al. Multi-national survey on the methods, efficacy, and safety on the post-approval clinical use of pulsed field ablation (MANIFEST-PF). Europace. 2022;24(8):1256–66.

5. Reddy VY, Ekanem E. Multi-national survey on the safety of the post-approval use of pulsed field ablation in 17000+ patients (MANIFEST-17K). Presented this at the AHA Late Breaking & Featured Science – Scientific Sessions November 11, 2023. Philadelphia, PA AHA. 2023.

6. Osmancik P, Bacova B, Hozman M, Pistkova J, Kunstatova V, Sochorova V, et al. Myocardial Damage, Inflammation, Coagulation, and Platelet Activity During Catheter Ablation Using Radiofrequency and Pulsed-Field Energy. JACC Clin Electrophysiol. 2023.

7. Balthazar T, Bennett J, Adriaenssens T. Hemolysis during short-term mechanical circulatory support: from pathophysiology to diagnosis and treatment. Expert Rev Med Devices. 2022;19(6):477–88.

8. Lusebrink E, Kellnar A, Krieg K, Binzenhofer L, Scherer C, Zimmer S, et al. Percutaneous Transvalvular Microaxial Flow Pump Support in Cardiology. Circulation. 2022;145(16):1254–84.

9. Huang JB, Wen ZK, Lu WJ, Lu CC, Tang XM. Diagnosis and Treatment of Mechanical Hemolysis after Mitral Repair in Adult. Heart Surg Forum. 2021;24(1):E165–E9.

10. Calvert PA, Northridge DB, Malik IS, Shapiro L, Ludman P, Qureshi SA, et al. Percutaneous Device Closure of Paravalvular Leak: Combined Experience From the United Kingdom and Ireland. Circulation. 2016;134(13):934–44.

11. Di Biase L, Diaz JC, Zhang XD, Romero J. Pulsed field catheter ablation in atrial fibrillation. Trends Cardiovasc Med. 2022;32(6):378–87.

12. Gass GV, Chernomordik LV, Margolis LB. Local deformation of human red blood cells in high frequency electric field. Biochim Biophys Acta. 1991;1093(2-3):162–7.

13. van Tilborg A, Dresselaars HF, Scheffer HJ, Nielsen K, Sietses C, van den Tol PM, et al. RF Ablation of Giant Hemangiomas Inducing Acute Renal Failure: A Report of Two Cases. Cardiovasc Intervent Radiol. 2016;39(11):1644–8.

14. Cannata A, Cantoni S, Sciortino A, Bruschi G, Russo CF. Mechanical Hemolysis Complicating Transcatheter Interventions for Valvular Heart Disease: JACC State-of-the-Art Review. J Am Coll Cardiol. 2021;77(18):2323–34.

15. Venier S, Vaxelaire N, Jacon P, Carabelli A, Desbiolles A, Garban F, et al. Severe acute kidney injury related to haemolysis after pulsed field ablation for atrial fibrillation. Europace. 2023;26(1).

